# A novel map of human p53 response elements uncovers evidence of selection pressures and variants similar to Li-Fraumeni Syndrome mutations

**DOI:** 10.1101/2022.09.28.22280453

**Authors:** Ping Zhang, Katherine Brown, David Barnes, Isaac Kitchen-Smith, David Sims, Adrienne Flanagan, Solange De Noon, Peter Van Loo, Steven Hargreaves, Will Cross, Nischalan Pillay, Mariam Jafri, Yvonne Wallis, Deniz Ucanok, Sotirios Ntaoulas, Markus G Manz, Toma Tebaldi, Alberto Inga, Genomics England Research Consortium, Lukasz F. Grochola, Gareth Bond

## Abstract

**Background:** The advent of functional genomic techniques and next generation sequencing has improved the characterization of the non-protein coding regions of the genome. However, the integration of these data into clinical practice is still in its infancy. Fifty percent of cancers mutate *TP53*, which promotes tumorigenesis, in part, by inhibiting its ability to bind to non-coding regions of the genome and function as a sequence-specific transcription factor. P53 is a tumour suppressor that inhibits cell survival through regulating transcription of anti-survival genes. However, p53 also regulates transcription of pro-survival genes and the target gene(s) responsible for p53 tumour suppression remains an open topic of research.

**Methods:** In this study, we integrate detailed genome-wide maps of p53 responsive elements (p53-RE), p53 occupancy, recently defined candidate cis-Regulatory Elements (cCREs) and whole genome sequencing for cancers to better define the regions of the genome that harbour functional p53 enhancers.

**Results:** We determine that p53-REs are more likely to be closer to the consensus binding site, to be evolutionarily conserved and to be occupied by p53 *in cellulo*, when they reside in regions of the genome that have been noted to have accessible DNA and a regulatory epigenomic mark in at least one human cell even without obvious p53 activation signals (cCRE p53-REs). We offer evidence that it is only in cCRE p53-REs, where multiple signs of differential natural selection between pro-survival and anti-survival target genes can be noted. Using whole genome sequences of 38,377 individuals, we go on to demonstrate that carriers of rare germline mutations in cCRE p53-REs can have similar traits to carriers of rare p53 coding mutations that cause the Li-Fraumeni cancer predisposition syndrome.

**Conclusions:** Together, these observations suggest that functional p53 enhancers are enriched in cCREs and that germline mutations in them have the potential to improve current cancer risk management and screening strategies.

## Background

Historically, the protein coding regions of both germline and cancer genomes have been the focus of much research which has defined many cancer driver mutations that have had substantial clinical impact in terms of managing cancer risk and developing precision therapies. Today, global projects like the Encyclopaedia of DNA Elements (ENCODE) have taken advantage of rapid increases in (epi-)genomic data to focus on the non-protein coding regions of the genome and define candidate cis-Regulatory elements (cCREs) in mice and man: regions of the genome with accessible DNA and regulatory epigenomic marks of transcriptional enhancers [1]. However, the integration of these data into clinical practice in oncology is still in its infancy. A crucial step in this process will be to determine which of these cCREs can be regulated by known oncogenes and tumour suppressors.

The tumour suppressor protein p53 regulates transcription of its target genes through its highly conserved, centrally located, sequence-specific DNA-binding domain (DBD). Indeed, there is overwhelming data that the ability of p53 to utilize this domain to bind DNA is crucial to its ability to act as a tumor suppressor. The p53 regulatory network is essential for maintaining genomic stability, preventing tumour formation and mediating the response to commonly used cancer therapies [2]. In addition, the p53 protein, a central node of this network, plays a key role in regulating other processes such as pigmentation, fecundity, cellular metabolism, mitochondrial respiration, stem cell maintenance and early embryonic development [3]. P53 mediates its action primarily through the induction of cell-cycle-arrest, apoptosis or senescence in response to a wide range of different cellular signals in a stimulus-and cell-type dependent manner. It determines these cellular fates predominantly by regulating the transcription of a wide range of target genes. Indeed, murine, cellular, and tumor model systems have clearly demonstrated that the ability of p53 to suppress tumor formation, in many tissue types, is dependent on its ability to regulate transcription [4]. However, which target genes and activities are essential for the impact of p53 on cancer remains actively debated [5-7]. No set(s) of target genes has conclusively been shown to mediate its tumour suppression. A necessary step in defining p53 target genes is to define its enhancer regions throughout the genome, which include its binding site(s).

The best evidence that sequence-specific DNA binding by p53 is required for its tumor suppressive activity comes from the study of both germline and tumor genetics of *TP53*. For example, 50% of human cancers carry somatic *TP53* mutations and over 80% of these are missense mutations spanning the DBD [8]. Under most conditions, p53 binds the consensus site as a homotetramer and subsequently recruits transcriptional co-factors to regulate transcription via an N-terminal transactivation domain [4]. The canonical p53 responsive element (p53-RE) is composed of two decamers of RRRCWWGYYY (R = purine; W = A or T; Y = pyrimidine), separated by a spacer of 0–13 nucleotides, leading to millions of putative p53-RE sites in the human genome. Indeed, evidence has been provided suggesting that p53 can employ an unsophisticated enhancer logic that is very uncommon among transcription factors. Specifically, data derived from human genome-wide studies suggests that p53 recognizes a core set of p53-REs regardless of cellular context, overriding variations in chromatin landscapes and nucleosome positioning, and without apparent need of auxiliary transcription factors [9].

Many of the same somatic DBD mutations can be found as inherited, cancer-causing mutations in exceptionally cancer-prone families that belong to the autosomal dominant cancer predisposition syndrome Li-Fraumeni (LFS) [10]. LFS is characterized by early cancer diagnosis and a high lifetime cancer risk, most notably for osteosarcoma, soft tissue sarcoma (STS) as well as other cancer types such as early-onset breast cancer, brain tumours, leukaemia, and adrenocortical carcinoma [11]. Clinical diagnostic criteria for the “classic” LFS kindred as well as the closely related cancer predisposition syndrome Li-Fraumeni-like (LFL) are well defined and include direct *TP53* mutation testing and genetic counselling [11]. To date, *TP53* mutation testing exclusively considers mutations in the coding regions of the gene, such as DBD mutations [12]. Germline mutations in *TP53* are identified in approximately 70% of families meeting the classic LFS and approximately 40% of families meeting the LFL diagnostic criteria, respectively [11]. However, even when the frequency of *de novo* mutations in *TP53* is considered, which is estimated to be between 7% and 20% [13], there is still a large group of individuals with a genetic predisposition to develop sarcoma for which a specific familial pathogenic/likely pathogenic (P/LP) *TP53* variant cannot currently be identified. Importantly, this shortcoming affects genetic counselling in clinical practice, since the management recommendations for this patient group differ from those with LFS (Version 2.2022, National Comprehensive Cancer Network® (NCCN®), nccn.org, accessed 10^th^ June 2022).

In addition to those rare germline mutations that can be found as inherited, cancer-causing mutations in LFS and LFL, common single-nucleotide polymorphisms (SNPs) in key bases of functional p53-REs have also been shown to influence the ability of p53 to regulate transcription and result in differences in cancer susceptibility [14, 15]. Indeed, one of the strongest cancer genome-wide association studies (GWAS) SNP resides in a key position of a functional p53-RE in the *KITLG* gene (rs4590952), a pro-survival p53 target gene [14]. P53 is best known for its inhibition of cell survival through regulating transcription of anti-survival genes. Upon severe DNA damage, p53 can facilitate apoptosis by inducing anti-survival genes e.g. *BAX, PUMA* and *NOXA*. However, p53 also has pro-survival target genes, like *KITLG*, and recent data from mouse models and human cancer cells suggest that cancers retaining wild type p53 benefit from the activation of these genes [16, 17]. For example, p53 can induce target genes for DNA repair and survival upon mild stresses (e.g., *CDKN1A* and *RRM2B*), as well as target genes to inhibit apoptosis, promote tumorigenesis and/or promote cell metabolism (e.g., *TRIAP1, TIGAR* and *ACAD11*) [18-20]. The observations described above suggest the possibility that p53-REs in the human genome could be under evolutionary selection pressure and that pro-survival and anti-survival p53-REs may have evolved differently. Indeed, multiple observations in the literature support this hypothesis. For example, it has been observed that p53-REs in apoptotic anti-survival genes reside in less evolutionarily conserved regions than REs in pro-survival cell cycle genes [21]. Additionally, it has been proposed that anti-survival p53-REs have evolved to have sequence elements that weaken p53’s ability to bind, thus creating weaker p53-REs [22, 23]. Differences in composition of the 4 most conserved bases in the RE, at positions 4, 7, 14 and 17 that make direct contact with the p53 protein have been proposed to underlie the differences in affinities between the REs in these two functional groups [24]. In more recent studies, these have been expanded to an additional six bases in the RE which are important for either the helical structure of the RE (G3, T8, A13, and C18) [25] or p53 target selectivity through its cofactor iASPP (C9 and G12) [26].

We and others have determined that in cells, there are over one million p53-binding sites in the genome. Moreover, 10% of these have been noted to be occupied in human cells [14, 27]. It is clear that not all of these are relevant to p53’s ability to function as a tumour suppressor. Indeed, as sequencing and characterisation of the non-coding regions of cancer patients and their cancers dramatically accelerate, it will become possible to identify whether or not deleterious cancer-causing mutations exist in specific p53-REs of certain target genes, thereby allowing for human genetics to help define the role of target gene regulation in p53 tumour suppression and further identify individuals at greater cancer risk. However, a crucial step in this process will be to improve our current definition of a functional p53-RE which will limit the currently overwhelmingly large multiple testing burden resulting from so many potential p53 binding elements in the genome. In this study, we attempt to do this by integrating detailed, up-to-date, genome-wide maps of p53 responsive elements (p53-RE), p53 occupancy, and the recently defined cCREs [1]. We utilize these data and present multiple lines of evidence that a subset of p53-REs (approximately 10%) are significantly enriched in multiple traits of functional p53 enhancers relative to the other 90% of all p53-REs in the human genome, including harbouring a p53-RE that is closer to the consensus binding site, evolutionarily conserved, and occupied by p53 *in cellulo*. We provide supportive evidence of the importance of cCREs in helping to define functional p53 binding sites in the genome, which allows insights into how these sites could help predict cellular phenotypes, guide genetic counselling, and improve current cancer risk management and screening strategies.

## Results

### 1. Identification of p53 binding sites in the human genome

To develop an up-to-date map of functional p53 binding sites genome-wide, we utilized newly available abundant genomic data and pattern search algorithms to generate detailed maps of p53-REs, p53 occupancy and genetic variations. First, we integrated p53 chromatin immunoprecipitation coupled with sequencing (ChIP-seq) data derived from 30 experiments utilizing nine different cell types including six different normal tissues and three different cancerous tissues [14, 28-39] (**Supplementary Table 1**). Cells were exposed to different methods to activate p53: 5-FU, Cisplatin, DMSO, Doxorubicin, Doxycycline, Etoposide, Nutlin-3a, retinoic acid, ionizing radiation or overexpression of RasG12V or E1A/RasG12V. The number of peaks called per dataset was highly variable (range: 1-26,331, median 1,395), as expected, given that these datasets were generated using diverse cell lines and with different, or no, p53 activating methods (**Supplementary Figure 1A**). Over 18 million base pairs, or 0.59% of the human genome, are covered by a ChIP-seq peak in at least one dataset, 0.21% of the genome was covered by a peak in two or more studies and 0.10% in three or more studies (**Supplementary Figure 1B**). The maximum coverage at any position in the genome was 26 datasets (**Supplementary Figure 1B**). For each coverage level, we merged covered regions of the genome from the different ChIP-seq datasets to generate sets of p53 bound intervals, resulting in intervals with a median length of 267 base pairs (range 1-11,546; **Supplementary Figure 1C-E**). In total, we identified 15,568 regions of the genome that were bound by p53 in at least one study.

**Table 1.** Classifications of p53 response elements depending on ChIP-Seq coverage level, the percentage of ChIP-Seq intervals at each coverage level which contain at least one putative RE as identified by p53 retriever, and the number of REs identified at each coverage level in the human genome. *C* represents the median number of ChIP-Seq datasets in which the RE was covered by a peak.

| Coverage Level | Classification | % Intervals at level *C* containing $\geq 1$ putative RE | #REs at level *C* |
| --- | --- | --- | --- |
| $C = 0$ | Unbound | NA | 1,455,529 |
| $1 \leq C < 3$ | Unlikely bound | 26.65% | 11,770 |
| $3 \leq C < 5$ | Lowly bound | 62.70% | 2,455 |
| $5 \leq C < 10$ | Moderately bound | 77.85% | 3,002 |
| $10 \leq C$ | Highly bound | 90.02% | 1,736 |

Next, we employed the p53retriever pattern search algorithm to map p53-REs genome-wide [27]. P53retriever is based on a set of manually curated rules, derived from a compendium of p53 transactivation data obtained using a yeast-based assay and ranks p53-REs according to the predicted transactivation potentials into five classes. In total, 1,474,492 putative p53REs were identified in the human genome using this algorithm. Of these, 83.52% scored one (unlikely), 12.25% scored two (poor), 3.21% scored three (slight), 0.93% scored four (moderate) and 0.08% scored five (high) (**Supplementary Figure 1F, Supplementary Table 2**). The ChIP-seq coverage level of each p53-RE identified with p53retriever was determined as the median (across the length of the p53-RE) number of ChIP-seq datasets in which the p53-RE was covered by a peak. 18,964 out of the 1,474,492 putative p53-REs were bound in at least one ChIP-seq dataset, with decreasing numbers of p53-REs at each subsequent coverage level (**Supplementary Figure 1G**). We categorized p53-REs of the genome as unbound, unlikely, lowly, moderately and highly bound if they were covered in 0, 1-2, 3-4, 5-9 and more than 10 datasets, respectively. As expected, the probability of binding increased together with the p53retriever score. Specifically, a large majority (84.00%) of unbound putative p53-REs had a p53retriever score of one and few (0.0024%) had a score of five (**Figure 1A)**. By contrast, only 16.01% of highly bound putative p53-REs had a p53retriever score of one and 23.79% had a score of five (**Figure 1A**). To quantify this overlap, a simulation analysis was performed using the Genomic Association Test (GAT) to measure the significance of the overlap between the intervals at these coverage levels and p53-REs with each score (**Figure 1B**) [40]. For the lowest scoring REs (score of 1) there is a small but significant enrichment in the overlap with p53 bound intervals at all coverage levels (FDR corrected p < 0.001; **Figure 1B**). This enrichment increases very rapidly with the score, confirming the utility of p53retriever in predicting the binding affinity of a p53-RE. With a score of 2 (“poor”), p53-REs have an approximately two-fold enrichment in highly p53 bound regions compared to the random expectation, which increases to a fold change of almost 2^12^ for the highest scoring p53-REs (scoring 5 or “high”) (**Figure 1B**, FDR corrected p < 0.001). As expected, we also found that higher proportions of highly bound and scored p53-REs had the consensus p53 binding site sequence (RRRCWWGYYY), relative to unbound or lowly scored p53-REs (**Figure 1C**). The p53 binding site sequence has primarily been determined through *in vitro* binding assays [5-7]. In line with the analysis above, p53retriever was more likely to predict regions bound by p53 in the ChIP-seq analysis with increasing coverage levels. Many intervals contained no p53-REs recognised by p53retriever at low coverage levels whereas the probability of containing a p53-RE was 62.70%, 77.85% and 90.02% if regions were covered in lowly, moderately and highly bound regions, respectively (**Figure 1D and Table 1**).

**Table 2:**
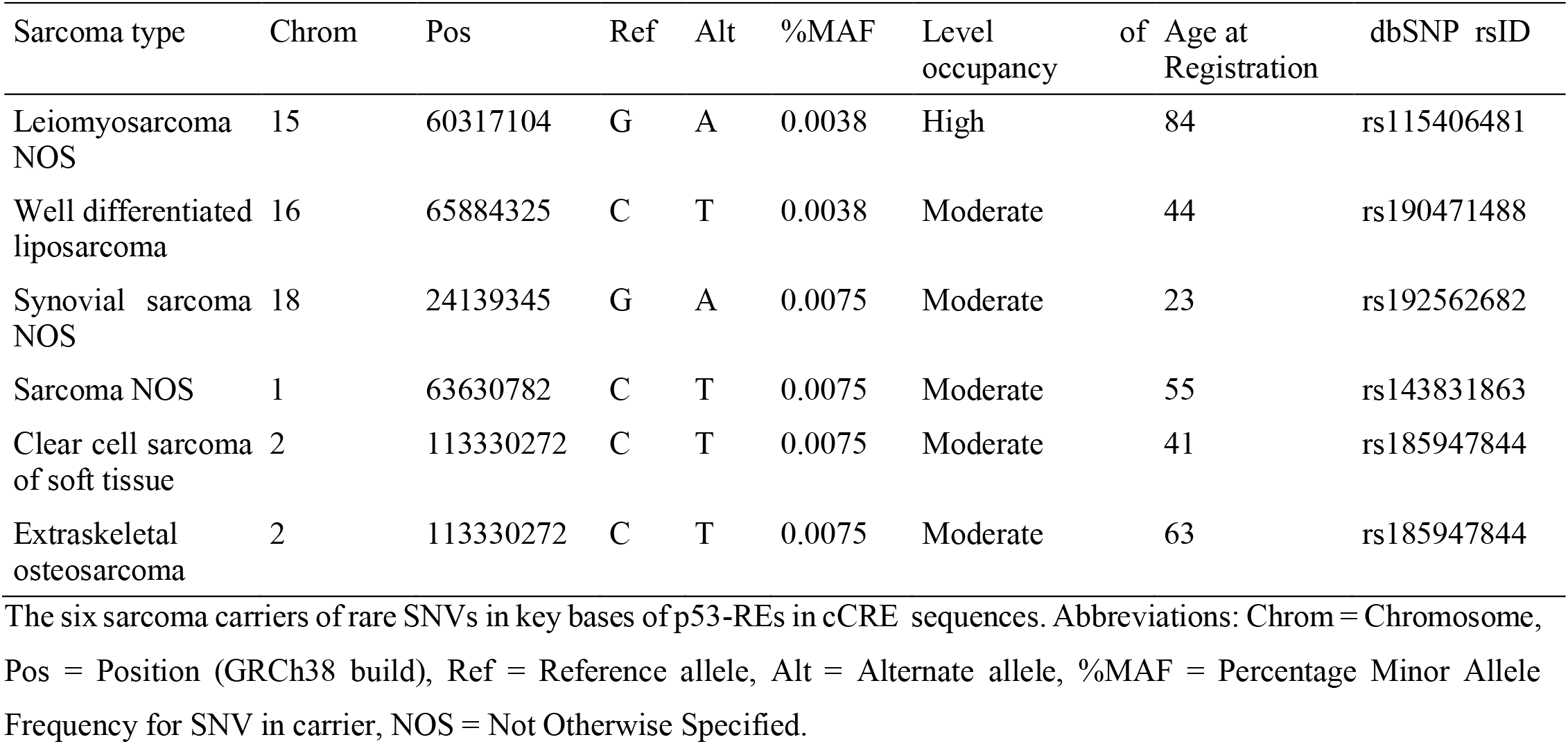
Sarcoma carriers of rare SNVs in key-base cCRE p53-REs.

**Figure 1:**
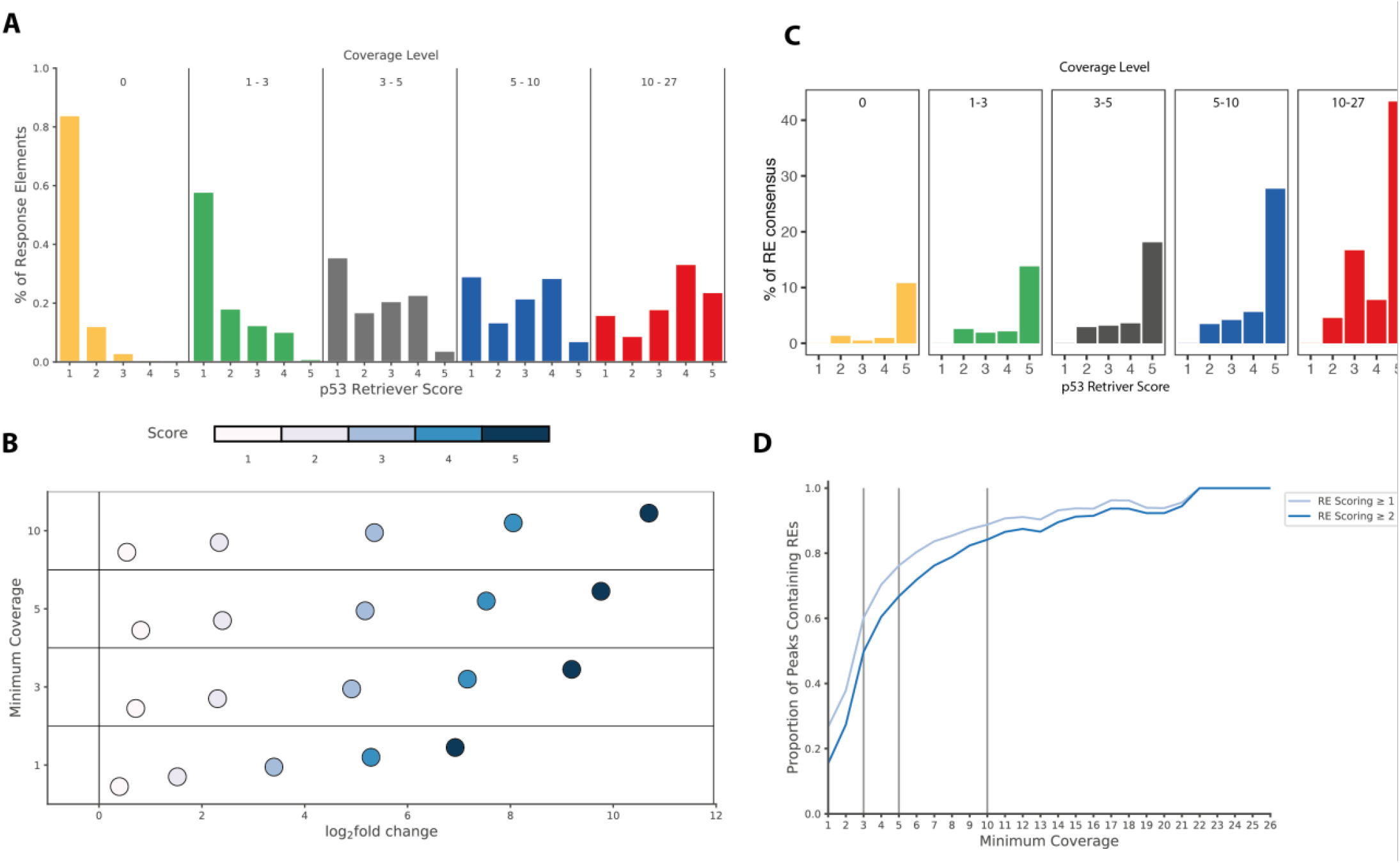
Identification of p53 binding sites in the human genome. (A) The percentage of p53-REs with each p53 retriever score for each level of ChIP-Seq coverage. (B) The log_2_ fold change compared to the random expectation in the overlap between intervals determined through ChIP-Seq coverage and p53 response elements with each possible p53 retriever score. All enrichments were significant with corrected p-values < 0.01. (C) Percentage of p53-RE consensus (RRRCWWGYYY) in each p53-RE coverage level and retriever score. (D) The proportion of ChIP-Seq intervals containing at least one p53-RE and containing at least one RE scoring ≥ 2 for intervals covered by peaks in increasing numbers of ChIP-Seq datasets.

**Supplementary Figure 1:**
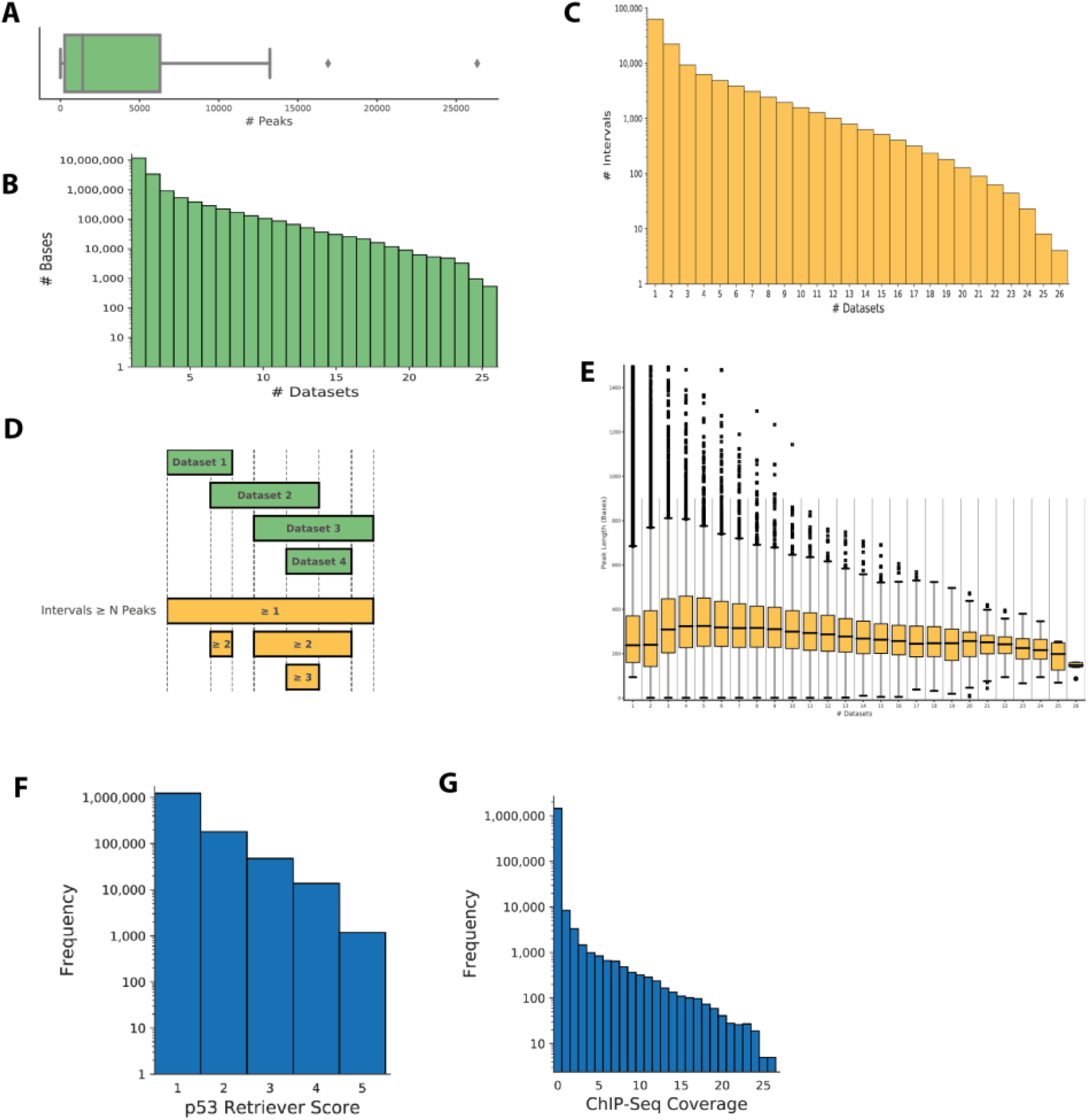
(A) The distribution of the number of peaks identified across the 30 ChIP-seq datasets. (B) The number of bases of the human genome covered by peaks in exactly N p53 binding datasets. (C) Bar plot showing the number of intervals in each set representing a minimum number of datasets. (D) The methodology used to identify intervals covered in at least N datasets. Green regions represent the peaks called in a series of (theoretical) datasets. Yellow regions represent the interval sets in which each region would appear. Each region of the genome can appear in multiple interval sets. (E) Box plot showing the distribution of interval lengths at each minimum coverage level. (F) Histogram showing the frequency of response elements with each possible p53 retriever score, where elements scoring 1 are classified as “unlikely” response elements, score 2 are “poor”, score 3 are “slight”, score 4 are “moderate” and score 5 are “high”. (G) Histogram showing the frequency of p53 response elements detected with the p53retriever algorithm covered by each number of ChIP-Seq dataset peaks.

### 2. Multiple response elements increase likelihood of p53 occupancy

Of the peaks containing at least one p53-RE, 17.02% (2,649 out of 15,568) contained two or more p53-REs (**Supplementary Table 3)**. We noted a significant trend wherebythe number of such peaks increased together with the coverage. Specifically, 13.33%, 27.00%, 33.60% and 40.21% of unlikely, lowly, moderately and highly bound peaks with p53-REs contained two or more p53-REs, respectively (p=5.09 × 10^−123^, Cochran-Armitage test; log_2_ fold change between highly bound and unlikely bound = 1.59, **Figure 2A** left panel). The difference between highly and lowly bound peaks was more pronounced with increasing p53retriever scores. Specifically, 23.91% of highly bound peaks containing p53-REs scoring two or more contained at least two such p53-REs, compared to only 3.67% of unlikely bound peaks (log_2_ fold change = 2.70; p = 6.15 × 10^−40^; Cochran-Armitage test; **Figure 2A** middle panel). Accordingly, 13.48% of such highly bound peaks, containing p53-REs scoring three or more, contained at least two p53-REs, compared to 1.56% of unlikely bound peaks (log_2_ fold change = 3.11; p = 5.82 × 10^−15^; Cochran-Armitage test; **Figure 2A** right panel). Only 24 peaks contained two or more p53-REs scoring four or more and no peaks contained two or more p53-REs scoring five, so these minimum scores were not analysed. These results indicate that the presence of multiple low to moderate scoring p53-REs can increase the probability of p53 binding.

**Figure 2:**
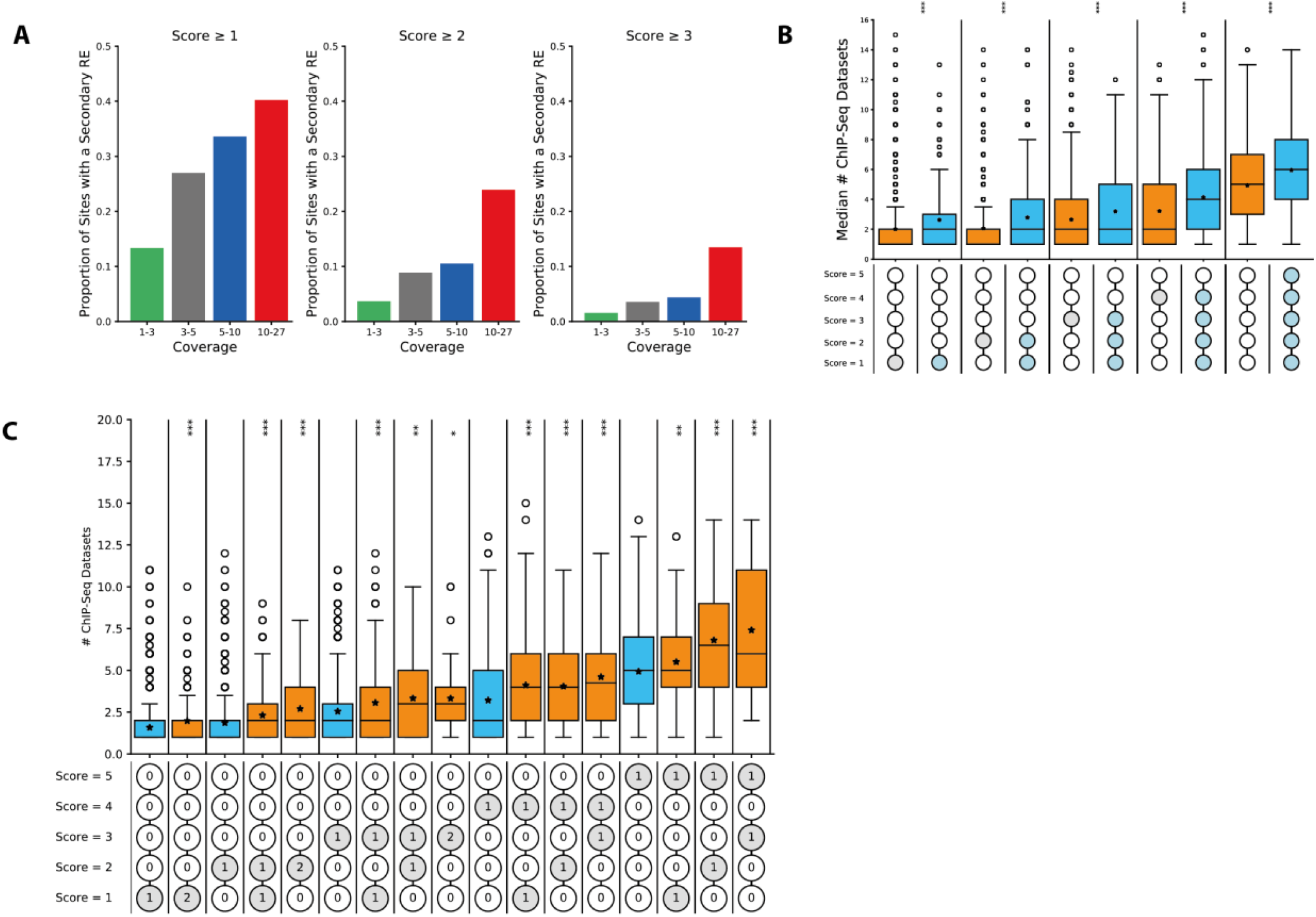
Multiple response elements increase likelihood of p53 occupancy. (A) The proportion of peaks containing ≥ 1 p53-RE (left), ≥1 p53-RE with a score of ≥ 2 (middle) and ≥ 1 p53-RE with a score of ≥ 3 which also contained a secondary p53-RE with the same score or higher for each coverage level. (B) Box plot showing the ChIP-Seq coverage of peaks containing a single p53-RE of a specific score (orange) vs peaks containing a secondary RE of this score or lower (blue). The bottom section of the graph shows the pattern of response elements corresponding to each box, with grey circles representing the score of the single REs and blue circles representing the potential scores of the secondary REs. (C) Box plot showing the ChIP-Seq coverage of peaks containing a single RE of a specific score (orange) vs peaks containing an RE with this score plus a specific secondary RE with this score or lower. The bottom section of the graph shows the pattern of response elements corresponding to each box, with grey circles representing the score of the REs in the comparison and numbers representing how many of this RE are present in the peak.

Given this result, we considered the possibility that either: (a) the probability of binding at a p53-RE is increased by the presence of a nearby p53-RE with the same score or lower, or (b) some combinations of p53-REs with particular scores increase the probability of binding. To test this hypothesis (a), all peaks containing exactly one (12,919 peaks) or exactly two p53-REs (2,206 peaks) were used. For each score *s* from one to five, the median number of ChIP-seq datasets covering the peak was compared between peaks with one p53-RE scoring *s* and peaks with one p53-RE scoring *s* and a “secondary” RE scoring ≤ *s* (**Figure 2B**). For all scores, there was a significant increase in binding when a secondary RE was present (s = 1, p = 1.25 × 10^−40^; s = 2, p = 1.79 × 10^−22^; s = 3, p = 1.93 × 10^−11^; s = 4, p = 2.65 × 10^−31^; s = 5 p = 1.52 × 10^−8^). To test hypothesis (b), the median number of ChIP-seq datasets covering all peaks containing exactly one p53-RE with a score *s* was compared to every possible combination of two p53-REs with scores ≤ *s* (**Figure 2C**). This analysis showed that all combinations of two p53-REs with scores ≤ *s* have significantly higher coverage than a single RE with score *s* (excluding combinations found in ≤ 20 peaks) (p < 0.05, Mann-Whitney U). Together, these results suggest that the presence of an additional p53-RE with a p53retriever score of two or higher, significantly increases the probability that p53 will bind at a particular position.

### 3. p53-REs in cCREs are more likely occupied by p53 and located in known target genes

As mentioned above, the ENCODE project has defined regions of the genome with accessible DNA (DNase hypersensitivity) and regulatory epigenomic marks (H3K4me3, H3K27ac, and/or CTCF-occupancy) in at least one human biosample as candidate cis-Regulatory elements (cCREs). To date, they have identified 1,063,878 cCREs in the human genome using 1,518 different biosamples [1]. We overlaid the defined cCREs and our defined p53-REs and determined that there are 151,800 p53-REs in cCREs (10.3% of all p53-REs, cCRE p53-REs) and 1,322,692 in non-cCRE regions genome-wide (89.7%, non-cCRE p53-REs, **Figure 3A**). As expected, 92% of the cCRE p53-REs are in defined enhancer regions. 79.1% are in distal enhancers (dELS; DNase hypersensitivity/H3K27ac) and 15.8% are proximal to gene promoters (pELS and PLS; DNase hypersensitivity/H3K27ac/H3K4me3). Interestingly, the cCRE p53-REs are more conserved throughout vertebrate evolution, as denoted by higher evolutionary conservation scores (average score 0.067 versus 0.017, p < 2.2e-16; **Figure 3B**). Surprisingly, the percentage of p53-REs in cCREs significantly increases as the RE’s predictive strength (Retriever score) increases, regardless of p53 occupancy levels: from 10% amongst REs with a score of 1 to 30% in REs scoring a 5 (p < 2.2e-16, **Figure 3C**). Moreover, the percentage of p53-REs in cCREs also significantly increases when p53 has been found to bind the RE *in cellulo*: from 10% in lowly bound REs to 60% in highly bound REs (p<2.2e-16, **Figure 3D**). Indeed, residing in a cCRE dramatically increases the likelihood of a p53-RE being occupied by p53. Specifically, 3.8%, 8.5%, 22.5%, 51.9%, or 93.7% of p53-REs with Retriever scores of 1, 2, 3, 4, or 5 are occupied by p53 in at least one of our datasets. In contrast, non-cCRE p53-REs are 10.1-fold less frequently occupied by p53 with Retriever scores of 1, 9.3-fold less with 2, 6.0-fold less with 3, 2.9-fold less with 4, and 1.5-fold less with 5 (p < 2.2e-16; **Figure 3E**).

**Figure 3.**
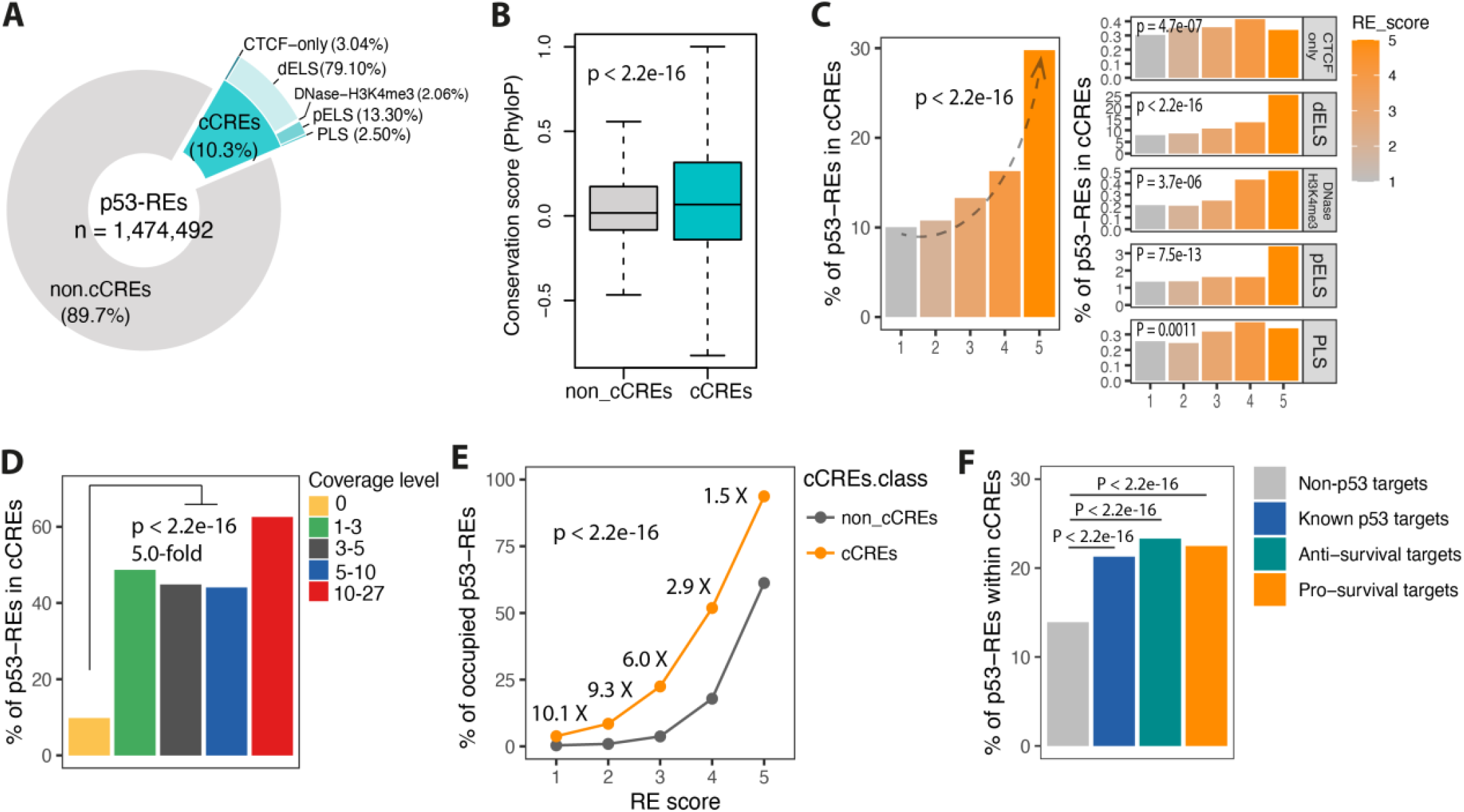
p53-REs in cCREs are more likely occupied by p53 and located in known target genes. (A) Fractions of p53-REs residing in indicated genomic regions. cCREs were defined by the ENCODE project as regions of the genome with accessible DNA (DNase hypersensitivity) and regulatory epigenomic marks (H3K4me3, H3K27ac, and/or CTCF-occupancy) [1]. (B). Box plot showing the average conservation scores of p53-REs that are in cCREs (cyan) or in non-cCREs (grey). P-value was calculated by Wilcoxon test. The outliers are not shown. (C). Bar plots showing the percentage of cCRE p53-REs with increasing p53-RE scores. P-values were calculated by Cochran-Armitage test. (D) The proportion of p53-REs in cCRE regions across the differing coverage levels. P-value was calculated by Fisher’s exact test. (E) Line plot showing the percentages of p53-occupied p53-REs in cCREs (orange) or not in cCREs (grey) regions across the different RE scores. P-values were calculated by Fisher’s exact test. (F) Bar plot of percentages of p53-REs in cCREs among different gene groups. REs were attributed to genes if they reside within a 10Kb window of an annotated gene boundary.

Collectively, these results support a model in which cCREs further define functional p53-REs, as cCRE p53-REsnot only have significantly higher levels of p53 occupancy, but also contain p53-REs that are more evolutionarily conserved and thus more likely to be functional (higher Retriever scores). To test this further, we reasoned that cCRE p53-REs should also be found more frequently in and around well-defined p53 target genes relative to other annotated genes. There are minimally 343 well-defined p53 target genes in the genome [41]. There are 14,251 p53-REs in and around (+/-10kb) the 343 target genes and 952,810 in and around the 58,411 other annotated genes. Only 13.9% of the p53-REs of the other annotated genes reside in cCRE regions, while significantly more, 21.3%, of the target gene p53-REs are in cCREs (OR = 1.7, p < 2.2e-16; **Figure 3F blue vs. grey bar**). Thus, cCRE p53-REs are found more frequently in proximity to well-defined p53 target genes, thereby lending further support to the model that cCRE p53-REs are more likely to be functional.

### 4. Signs of differing selection pressures on anti-survival and pro-survival p53-REs are limited to cCREs

In this study so far, we have taken advantage of newly generated functional genomic data to see if we can improve our current definitions of a functional p53-RE with the aim of reducing the large multiple testing burden by excluding many non-/less-functional p53 binding sites throughout the genome. We have provided multiple lines of evidence supporting the model that cCRE p53-REs are more likely to be functional. As mentioned above, previous publications have noted significant differences between p53-REs in anti-survival and pro-survival target genes. We therefore reasoned that if cCREs are more likely to be functional, then we should expect the differences between p53-REs in anti-and pro-survival target genes to be most apparent in cCREs p53-REs. One of the most studied differences between the REs of these two different groups of target genes is their predicted strength. Specifically, it has been proposed that anti-survival genes have evolved weaker p53 response elements to ensure the requirement of additional factors to recruit p53 and up-regulate these irreversible stress responses [22, 23]. Therefore, we wanted to explore in our dataset if there was any support for differential selection pressures on functional p53-REs depending on whether or not they reside in anti-or pro-survival target genes. To test this, we first annotated the 343 known p53 target genes into three classes (anti-survival, pro-survival and other) based on their reported functions in model systems [41] (**Supplementary Figure 2A** and **Supplementary Table 4**). Anti-survival target genes include genes responsible for pro-apoptotic, anti-proliferative and/or pro-senescence activities, while pro-survival target genes include genes responsible for anti-apoptotic, pro-proliferative and/or pro-metabolic activities (**Supplementary Figure 2B**). Amongst the 343 well-defined p53 target genes [41], 130 genes have at least one occupied p53-RE in only cCRE regions (37.9%), with 33 genes having two, 18 having three, and 11 having four or more. Of the 36 genes without a p53-RE in a cCRE, 28 have at least one RE in only non-cCRE regions (77.8%), with four genes having two, and four having three. 133 genes (38.8%) have at least one RE in both cCRE and cCRE regions. 44 genes (12.8%) genes do not have any occupied p53-REs. Interestingly, we found that occupied cCRE p53-REs (≥1 ChIPseq) in anti-survival genes are significantly weaker (lower average Retriever Scores) than cCRE p53-REs in pro-survival genes (median score 1 for anti-survival p53-REs vs. score 2 for pro-survival REs. P = 0.020; **Supplementary Figure 2C**). In contrast, no differences were seen in occupied non-cCRE p53-REs (**Supplementary Figure 2D**), lending support to the model that cCRE p53-REs are more likely to be functional.

We next wanted to extend these comparisons of cCRE p53-REs and non-p53-REs the previously published observation that p53-REs in apoptotic anti-survival genes reside in less evolutionarily conserved regions than REs in pro-survival cell cycle genes [21]. To do this, we calculated median conservation scores (PhyloP) for the p53-REs in either cCREs or non-cCREs at the various levels of p53 occupancy for either pro-survival (**Figure 4A** left panel) or anti-survival (**Figure 4A** right panel) target genes. It becomes clear that the medians of the average conservation scores of pro-survival cCRE p53-REs are significantly greater than p53-REs in non-cCRE regions (**Figure 4A** left panel). No such differences were observed with anti-survival p53-REs (**Figure 4A** right panel). These highly conserved pro-survival p53-REs were found in cell cycle genes such as *CDKN1A* and *PLK2* that were noted to have conserved REs [21] as well as *ARHGEF3, RPS19, RHOC, LIMK2* and *BCL2L1* promoting cell survival via diverse pathways (**Supplementary Table 5**). It is important to note that no differences in conservations scores between anti-survival and pro-survival genes were found in other genic regions, such as UTRs, stop and start codons, and coding regions (**Supplementary Figure 2E**). In summary, the previously noted differences in conservation of p53-REs in pro-survival and anti-survival genes come from the cCRE p53-REs, as no differences were seen in non-cCRE p53-REs, lending further support to the model that p53-REs in cCREs are more likely to be functional.

**Figure 4:**
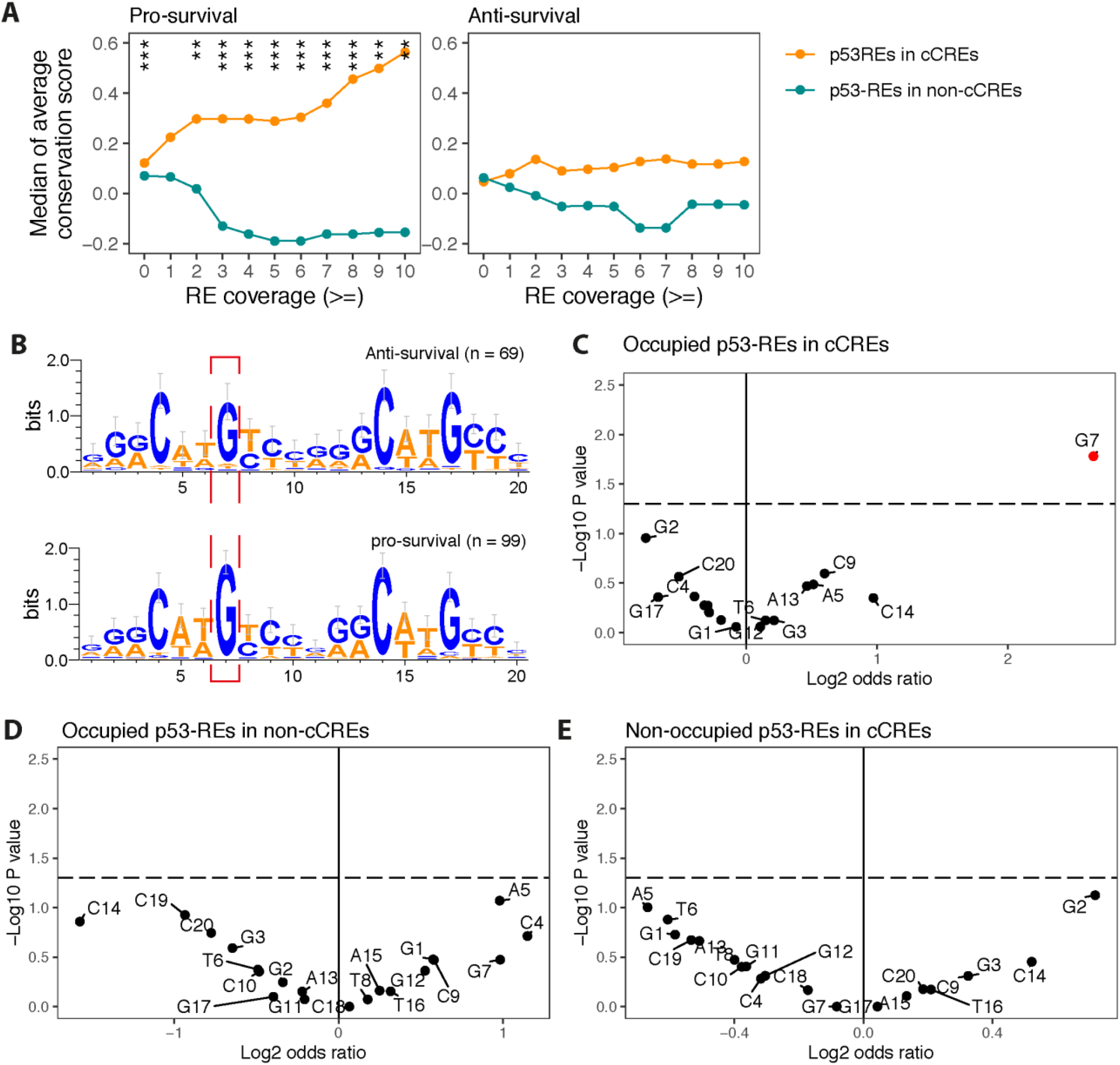
Pro-survival p53-REs are stronger and more likely to undergo genetic selection. (A). Line plots showing the Retriever scores of the p53-RE in cCRE (orange) or non-cCRE (cyan) regions that are proximal to the same set of genes (left panel: pro-survival; right: anti-survival). (B). Sequence logos showing the nucleotide distributions of the 20-bp consensus of the occupied pro-survival (lower) or anti-survival p53-REs (upper) in cCRE regions. Spacers between half sites of the REs were removed. Logos were generated using WebLogo (http://weblogo.berkeley.edu/). (C-E). Scatterplot showing the enrichment of each nucleotide of the 20-bp p53-binding motif on the *x* axis (Log2 odds ratio) and the corresponding *P*-value on the *y* axis (−log_10_ scale) found in the pro-survival group vs. anti-survival group. P-values and odds ratios were calculated by two-tailed Fisher’s exact test. The horizontal dashed lines correspond to a P-value of 0.05.

Our work thus-far suggests that differences in p53-REs predicted strengths between these two functional groups are primarily found in cCRE p53-REs. We therefore wanted to explore if the previously noted differences in specific bases that make direct contact with the p53 protein and/or mediate RE structure and selectivity, are also found when we compare the REs in cCREs. Indeed, when we take the occupied (≥1 ChIP-seq) p53-REs in cCREs with the highest RE score for each gene and compare the REs between the pro-survival and anti-survival genes, we note a significant enrichment of the G7 (contact/key base) in pro-survival REs relative to anti-survival REs (**Figure 4B-C**). Specifically, we determined that 97 of the 99 pro-survival p53-REs have a G allele at the 7th position (key base) compared to 61 out of the 69 anti-survival REs (P= 0.017, OR = 6.3; **Figure 4B**) providing further evidence that functional pro-survival p53-REs are more likely to have the key contact bases, and hence stronger p53 affinities. Consistent with our previous findings, no significant enrichments were seen in any of these 10 bases of the non-cCREs p53-REs or in the non-occupied cCRE p53-REs (**Figure 4D-E)**, thereby lending further support to the model that cCRE p53-REs are more likely to be functional.

**Supplementary figure 2.**
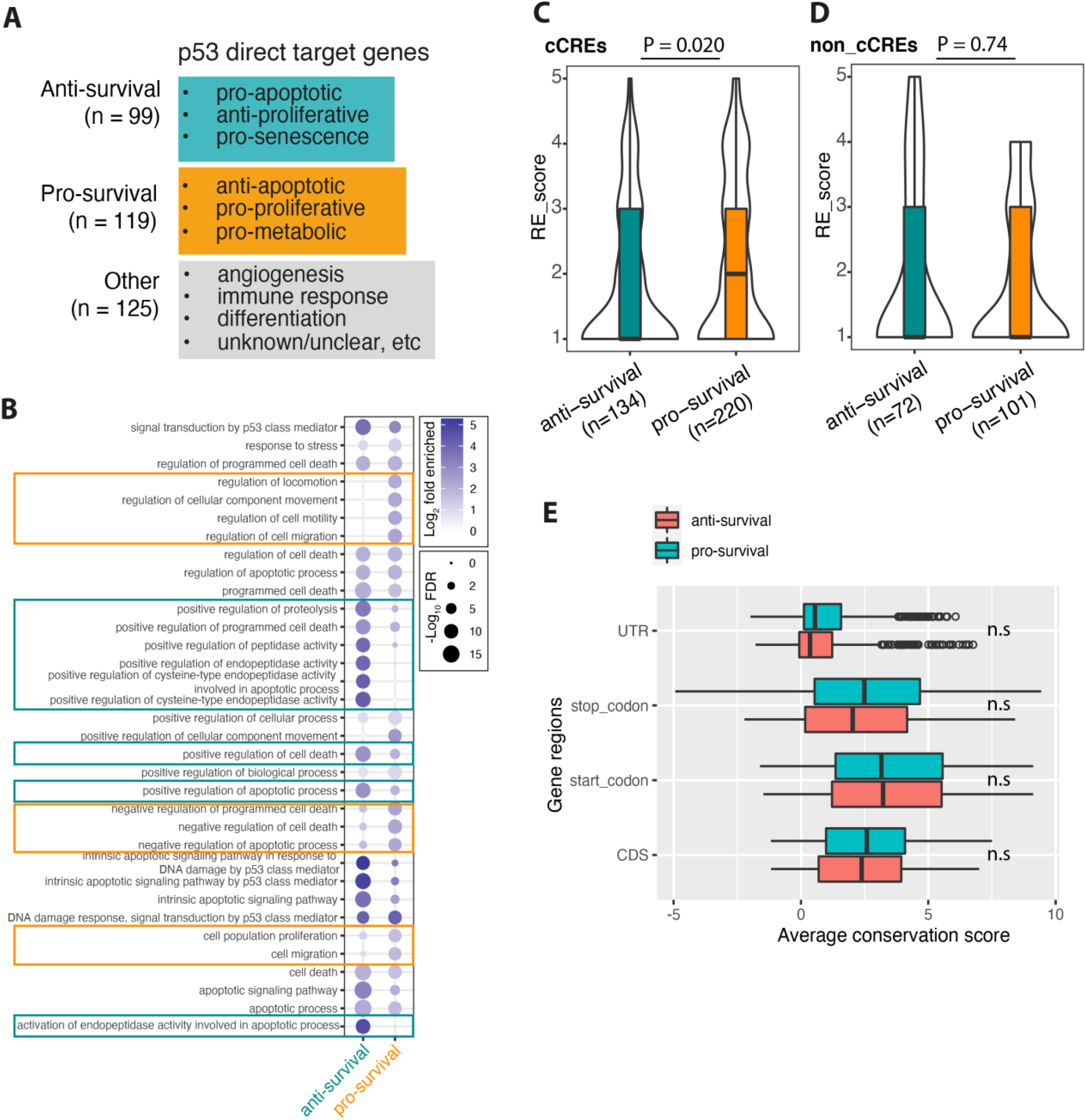
(A). Functional classification for 343 known p53 direct target genes and the number of genes in each class (see also **Supplementary Table 4**). (B). Top 10 significantly enriched Gene Ontology (GO) terms based on enrichment analysis of anti-survival or pro-survival p53 direct target genes using R package topGO. The pro-survival and anti-survival pathways were highlighted in orange and cyan boxes, respectively. (C-D). Box plot showing the retriever scores of occupied anti-survival (blue) and pro-survival (orange) p53-REs in cCREs (C) or non-cCRE (D) regions. P-value was calculated by a Wilcoxon test. (E). Box plot showing the average conservation scores between pro-survival (blue) and anti-survival genes (red) across different regions of gene body including UTR (untranslated region), start codon, stop codon and CDS (coding DNA sequence). P-values were calculated by Wilcoxon tests. n.s: not significant.

### 5. Individuals with germline mutations in cCRE p53-REs show similarities to LFS mutation carriers in the Genomics England Cohort

Thus far, all our comparisons of p53-REs in either cCREs or non-cCREs, support the model that cCRE p53-REs are more likely to be functional, as well as underlining the biological importance of p53-REs when they reside in regions of the genome that have been noted to have accessible DNA and a regulatory epigenomic mark in at least one human cell, even without obvious p53 activation signals. Therefore, we next wanted to extend these comparisons to human genetic health data and explore whether or not rare mutations in any type of p53-RE could lead to increased cancer risk. As mentioned above, the best evidence that the binding of p53 to its RE is crucial in its tumour suppression comes from the study of both germline and tumour genetics of *TP53*. 50% of human cancers carry somatic mutations in the *TP53* gene, over 80% of which are missense mutations spanning the DBD [8], and many of the same somatic DBD mutations can also be found as inherited, cancer-causing mutations in exceptionally cancer-prone families with Li-Fraumeni syndrome (LFS) [10]. Currently, there are 130 distinct known pathogenic and/or likely pathogenic (P/LP) mutations in the *TP53* gene associated with LFS in ClinVar, one of the most utilized aggregators for genomic variants associated with human diseases [42] (**Supplementary Table 6**). Only recently, with the advent of whole genome sequencing of participants in large health studies is it possible to explore whether rare mutations in p53-REs also lead to increased cancer risk and help explain part of the missing heritability in genetic predisposition syndromes such as LFS. Thus, we wanted to apply our novel maps of p53-REs to the genomes of the participants of the Genomics England project to explore this possibility.

In total, we studied the germline, whole genomes of 13,291 individuals of European ancestry who had been diagnosed with cancer (cancer participants), and 25,086 individuals of European ancestry who were relatives of a proband recruited with a rare disease not directly associated with increased cancer risk (controls, **Material and Methods**). We reasoned that if germline mutations in p53-REs lead to an increased risk of cancer, then the cancer participants should be enriched in these carriers relative to the controls. To explore this possibility, we first wanted to ensure that this is the case for known cancer causing mutations in the *TP53* gene itself, and/or other known tumor suppressors (*BRCA1, BRCA2, MSH6*, and *PALB2*). We wanted to directly compare the possible effects with the rare P/LP mutations of *TP53*, thus we restricted our subsequent analyses to those variants, which had minor allele frequencies in our controls group at or below the most frequent known *TP53* LFS P/LP (rs28934576 ≤0.02%; **Supplementary Table 6**). In total, we identified 12, 47, 107, 39, and 21 carriers of P/LPs based on ClinVar [42] (see Methods) in *TP53, BRCA1, BRCA2, MSH6*, and *PALB2*, respectively, amongst both the cancer participants and the controls. It is important to note that we also identified P/LPs in these genes at variant allele frequencies (VAFs) outside the typical VAF range (35% to 65%) used by clinical laboratories to call germline heterozygous variants. As expected from previous studies and its role in haematopoiesis [43], *TP53* had a high proportion of these potentially mosaic carriers: 12 carriers of P/LPs within the germline range and 12 carriers outside the range (50%, **Figure 5A**). It is notable that this level of potentially mosaic P/LP carriers is significantly higher than the levels found in the four other tumour suppressors (OR = 12.3, 95% CI 4.36 to 35.5; p = 5.11 × 10^−7^, **Figure 5A**). Moreover, *TP53* is the only gene amongst the five tumor suppressor genes wherein the levels of potentially mosaic mutation carriers are significantly higher when analysing P/LPs, than the levels of carriers of known benign or likely benign mutations in the same gene (B/LBs in ClinVar, Methods, p = 9.63 × 10^−6^, **Figure 5A**). In terms of enrichment of germline P/LP carriers amongst the cancer patients relative to the controls, all five tumour suppressor genes demonstrated significant enrichment in cancer types the protein products are known to suppress (**Figure 5B**): *TP53* in sarcoma, *BRCA1* and *BRCA2* in breast and ovarian cancers, *MSH6* in colorectal and endometrial cancers, and *PALB2* in breast cancer. Moreover, these significant enrichments of P/LP carriers in these cancer types were not found for B/LB carriers for any of these gene/cancer type pairs, lending further confidence for the specificity and biological relevance of these associations in the Genomics England cohort (**Figure 5C**).

**Figure 5:**
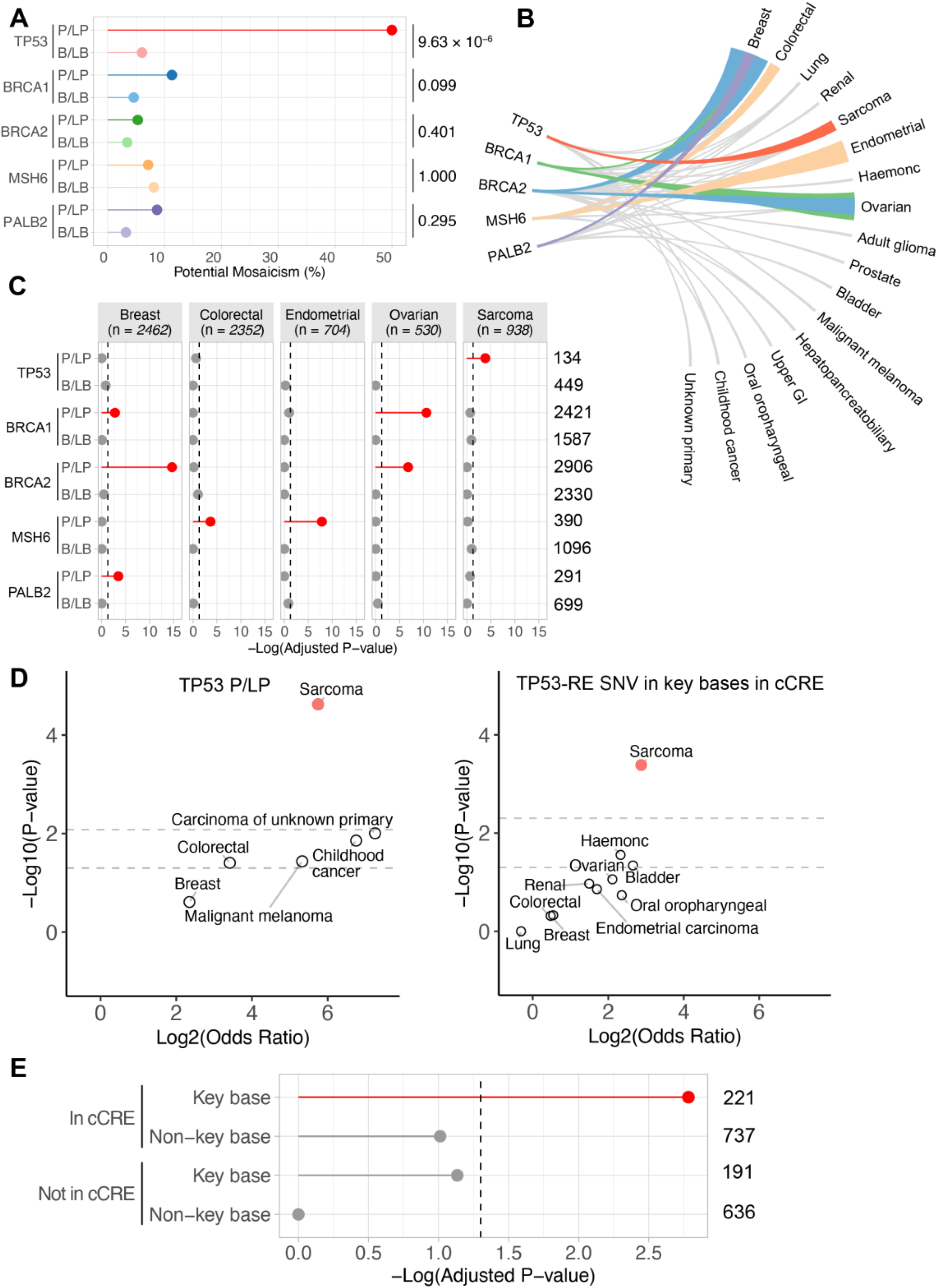
Rare germline SNVs in p53-REs are enriched in sarcoma participants in the Genomics England Cohort in a similar manner to LFS mutations (A) Lollipop chart showing the percentage of P/LPs or B/LBsvariants that were most likely to be caused by somatic mosaicism for 5 tumour suppressor (TS) genes in the European GEL cancer and control cohorts. Percentages were calculated for each gene for the combined cancer and control cohorts. Values to the right of the figure are raw p-values for Fisher’s test of enrichment comparing likely mosaicism in P/LPs to likely mosaicism in B/LBs. (B) Widths of ribbons adjacent to names of cancer types are proportional to –log10(adjusted p-value) from Fisher’s tests of enrichment of TS P/LPs in cancer types. Links in grey indicate that P/LPs were found in a cancer type but were not significantly enriched. (C) P/LPs in the 5 TS genes, but not B/LBs, were significantly enriched in specific cancer types. Dashed lines indicate the threshold of significance for p-values from Fisher’s tests of enrichment after adjustment for FDR. Lollipops are grey if not significant and red if significant. Absence of a lollipop in a cancer type indicates that a P/LP or B/LB for that TS gene was not detected. Numbers, n, listed under the names of cancer types are the sizes of each cohort in GEL. Values to the right of the plot are the numbers of rare (minor allele frequency < 0.0037%) P/LPs or B/LBs, respectively, for each gene in the high-confidence ClinVar sets. (D, Left) Germline *TP53* P/LPs are highly-significantly enriched in sarcoma but no other cancer types. Dashed lines indicate the thresholds for significance for p-values obtained from Fisher’s tests for enrichment before (lower) and after (upper) adjustment for multiple hypothesis testing. (Right) Germline SNVs in key bases of cCRE TP53-REs are significantly enriched in sarcoma but no other cancer type. Dashed lines indicate the same thresholds for significance as in the left scatterplot. (E) In sarcoma, SNVs in p53-REs are highly-significantly enriched in key bases in cCRE sequences (adjusted p-value = 0.0016) but not in non-key bases in cCRE sequences or in non-cCRE sequences. The dashed line indicates the threshold for significance for p-values from Fisher’s tests of enrichment adjusted for multiple hypothesis testing. Lollipops in red are significant whereas those in grey are not. The values to the right of the plot are the numbers of SNVs in cCRE or non-cCRE sequences and in key bases or non-key bases, respectively, in the sets that were used to interrogate germline variants called in the GEL aggV2 gVCF aggregation.

To determine if any similar associations could be found in the cancer participants with deleterious mutations in functional p53-REs, we first overlaid our maps of p53-REs found occupied by p53 in at least five ChIP-seq datasets, with the human SNP database of germline genetic variants (dbSNP). We wanted to directly compare the possible effects of these single nucleotide variants (SNVs) with the rare P/LP mutations of *TP53*, thus we restricted our subsequent analyses to those variants, which had minor allele frequencies in our controls group at or below the most frequent *TP53* LFS P/LPs (≤ 0.02%, rs28934576). We identified a total of 1,785 carriers with rare p53-RE SNVs: 221 carriers had an SNV in a cCRE p53-RE and in a key contact base, 737 in a cCRE p53-RE in a non-key contact base, 191 in a non-cCRE p53-RE and in a key contact base, and 636 in a non-cCRE p53-RE in a non-key contact base. In total, we found 426 carriers of p53-RE SNVs in the Genomics England (GEL) cancer and control cohorts, within the typical VAF range (35% to 65%) used by clinical laboratories to call germline heterozygous variants. In contrast to the P/LP calls, there was no evidence of any significant enrichment of carriers with potentially mosaic p53-RE SNVs, regardless of their genomic or RE locations (OR = 0.668, adjusted p-value (FDR) = 0.320).

When we calculated the potential enrichment of carriers of p53-RE SNVs in the cancer participants relative to the controls, we found very similar associations to those found with *TP53* P/LPs (**Figure 5D**). Specifically, we noted that carriers with SNVs in key bases of cCRE p53-REs were over seven-fold enriched amongst sarcoma patients (in six, out of a total of 938 sarcoma patients relative to 22//25,086 controls, OR = 7.33, adjusted p-value (FDR) = 0.0045, **Figure 5D**, right panel). This enrichment was not observed in other cancer types and mirrors the enrichment pattern seen in carriers of TP53 P/LPs (Sarcoma OR = 53.7, adjusted p-value (FDR) = 1.42 × 10^−4^, **Figure 5D**, left panel). When we expanded these analyses to SNVs found in either non-cCRE regions or other bases of the RE, we note that only carriers of SNVs in key p53 contact bases of p53-REs in cCRE regions were significantly enriched in the sarcoma patients (n = 10) relative to the controls (n = 49), thereby lending further support to the p53-dependency of these associations and the importance of cCREs in p53 enhancers (key base in cCRE, OR = 7.33, adjusted p-value (FDR) = 0.0016). (**Figure 5E, Table 2**). It is important to note that all SNVs in key bases of the cCRE p53-REs in the sarcoma patients are predicted to weaken p53 binding (**Table 2**).

## Discussion

In this study, we have aimed to take advantage of newly generated functional genomic data to improve our current definitions of functional p53 response elements. We generated a comprehensive genome-wide map of p53 response elements by incorporating sequence analyses and detailed genomic functional information on p53 binding sites. In our analyses, we found approximately 1.5 million putative p53-REs genome-wide using the p53retriever algorithm which ranks REs according to the predicted transactivation potentials (scoring 1 to 5) derived from data generated in yeast-based assays. As expected, a RE with a higher score was closer to the consensus p53 binding site (**Figure 1C**) and was more likely to be bound by p53 in at least one of the ChIP-seq datasets we processed (**Supplementary Figure 1G**), which were derived from 30 experiments utilizing nine different cell types and eleven different p53-activating methods. We went on to show that approximately 10% of the putative p53-REs reside in cCREs: in regions of the genome that have been noted to have accessible DNA and a regulatory epigenomic mark in at least one human cell even without obvious p53 activation signals (**Figure 3A**). p53 has been shown to be able to bind to its RE when found in both closed and open chromatin in multiple experimental settings and indeed p53’s role as a pioneer factor has been discussed [9, 44]. Our results suggest that, in most cases, functional p53 enhancers have evolved in regions that are more likely to be in open chromatin even before p53 is activated. Specifically, we found that cCRE p53-REs were more likely to have greater transactivation lpotential, be evolutionary conserved, bound by p53 *in cellulo*, and reside in known p53 target genes, suggesting that this group of p53-REs are enriched in functional p53 enhancers (**Figure 3**). In further support of this, we demonstrate that the previously reported signs of differential evolutionary selection pressure between p53-REs found in either anti-survival or pro-survival target genes can only be found amongst cCRE p53-REs [21, 23]. Specifically, only in p53-occupied cCREs p53REs are anti-survival target genes less conserved, weaker, and less likely to have a key contact base in the consensus-binding site (**Figure 4**).

Together, these data not only suggest that predicted p53-REs in cCREs are more enriched in functional p53 enhancers, but the results of our comparisons of p53 bound cCRE p53REs also make clear that, on average, p53-REs in pro-survival target genes have greater average transactivation potential (higher Retriever scores (**Supplementary Figure 2C-D**) and are more conserved (**Figure 4A**). We identified a total of 220 occupied pro-survival p53-REs in cCREs (**Supplementary Table 5**), 31% (62 out of 220) of which are highly occupied (ChIPseq datasets ≥10). These highly-occupied p53-REs reside in 50 unique genes that are known to be up-regulated after p53 activating cancer treatments, including oncogenes such as *MDM2* and *NOTCH1* [45], as well as genes that attenuate cellular responses to DNA-damaging agents and promote cell survival via diverse mechanisms. For example, *TRIM32* forms a negative feed-back loop with p53 to promote tumorigenesis [46], *RHOC, LIMK2* [47] and *KITLG* [17] protect cells from p53-mediated DNA damage response and apoptosis via downstream pro-survival pathways, and *TNFRSF10C* [48], *TNFRSF10D* [49] and *TNFAIP8* [50] antagonise TRAIL-induced cell apoptosis and/or promote inflammation. This suggests that the inhibition of the pro-survival/pro-tumor signalling mediated by these p53 target genes may synergise with many anti-cancer treatments that activate p53, as has been demonstrated in our recent study for the p53 activated pro-survival signal mediated by the cKIT signalling pathway [17]. Interestingly, 54% (27 out of 50) of the protein products of these transcripts could be targeted either directly or reside in druggable pathways [51] (**Supplementary Table 5**).

The missing heritability in LFS and related syndromes remains an area heavily studied in the field, but no conclusive results have yet to be obtained. It has been estimated that 20% of families with a diagnosis of LFS or LFS-related syndromes have no known pathogenic TP53 mutation or other cancer-associated mutations [52, 53]. Our observations lend strong support to the model that there is a subset of predicted p53-REs (approximately 10%) that are significantly enriched in multiple traits of functional p53 enhancers relative to the other 90% of all predicted p53-REs. These p53-REs can be a focus of future genetic and epigenetic studies of p53 enhancers, which would limit the dramatically large multiple testing burden with so many potential functional p53 binding elements in the genome (over 1.5 million). Encouragement of such studies comes from our results that rare single nucleotide germline mutation carriers in key bases of cCRE p53-REs are 7.3-fold enriched amongst sarcoma patients of the Genomics England cohort compared to our control group (p=4.12 × 10^−4^, adjusted p= 0.0016, **Figure 5**). The fact that carriers of both P/LPs in TP53 and SNVs in key p53-contact bases of cCRE p53-REs are only significantly enriched in sarcoma patients out of 23 other cancer types is striking and underlines the importance of p53 tumour suppression of this cancer type. The lifetime increased risk for carriers of *TP53* pathogenic mutation carriers for developing sarcomas is well known. Indeed, in almost all subtypes of LFS, a sarcoma diagnosis in a family can be a defining hallmark [54].

In the Genomics England cohort 103 different sarcoma subtypes were represented, and the patients were recruited from 11 different hospitals in England (**Supplementary Table 7**) with a median average age at registration of 63 years. None of the sarcoma patients with germline TP53 P/LPs where previously known to be p53 P/LP carriers until their genomes were sequenced by Genomics England (Solange De Noon, personal communication). Those four p53 P/LP carriers amongst the sarcoma cohort were diagnosed with three different sarcoma types: pleomorphic liposarcoma (x2), leiomyosarcoma and periosteal osteosarcoma and recruited at a mean average age of 60.8 years and a median average age of 67. These sarcoma types are known to be associated with germline p53 mutations. Indeed, they can be found amongst p53-mutation carriers in the IARC database R20 (July 2019) [55], which contains *TP53* variant data compiled from those published in the literature since 1989 or those present in other public databases. Of note, two of four sarcoma patients carried the very same rare gain-of-function TP53 P/LP rs11540652. The six carriers of SNVs in key contact bases of cCRE p53-REs with a sarcoma diagnosis were diagnosed with five different, defined sarcoma types: leiomyosarcoma, well differentiated liposarcoma, synovial sarcoma, clear cell sarcoma of soft tissue and extraskeletal osteosarcoma (**Table 2**). The diagnosis for the sixth carrier was not well-defined: sarcoma NOS (Not Otherwise Specified, **Table 2**). The mean average age at registration for the six carriers was 52 years and the median was 49 years (**Table 2**). Four of these well-defined sarcoma subtypes are found amongst p53-mutation carriers in the IARC database: 72 leiomyosarcoma, 33 liposarcoma, 1 synovial sarcoma and 280 osteosarcoma patients [55]. One of the well-defined sarcoma subtypes, clear cell sarcoma of soft tissue, is not present in IARC. Thus four out of the five sarcoma types diagnosed amongst the six carriers of SNVs in key contact bases of cCRE p53-REs are already known to be associated with mutations attenuating p53 pathway activity. Although the rare SNVs in these sarcoma patients reside in p53-occupied REs, the REs are not in or near known p53 target genes. However, they are predicted to decrease p53’s affinity to binding sites (**Table 2**), thus suggesting a possible attenuation of p53 signalling mediated by potentially novel target genes. If validated in additional cohorts, our results suggest that the non-coding regions of the genome might harbour rare mutations that can affect p53-signalling in a way that attenuates tumour suppression, which could help identify some of the missing heritability in LFS or other related syndromes and/or other p53-surveilled cancer types. The identified SNVs have the potential to be utilised in a clinical setting by genetic counsellors and further improve current sarcoma risk management and screening strategies.

## Methods

### ChIP-Seq analysis

Reads from 30 ChIP-seq datasets (listed in **Supplementary Table** 1) were downloaded from the Sequence Read Archive (SRA) (https://www.ncbi.nlm.nih.gov/sra). All datasets consisted of single ended Illumina reads. If multiple conditions were used in the same experiment these were incorporated as separate datasets. Reads were trimmed using Trimmomatic version 0.32 [56] and bases with leading or trailing quality less than 3, across a 4 base sliding window with quality less than 15 were trimmed, as were Illumina adaptors. Reads with greater than 24 bases remaining were retained. Reads were mapped to hg38 using the BWA-mem alignment algorithm version 0.7.12 [57]. The resulting BAM files were filtered to remove unmapped reads, duplicate reads (as identified with Picard MarkDuplicates 2.8.3 (http://broadinstitute.github.io/picard/)) and reads with a mapping quality score less than 10. Peaks were called using macs2 (version 2.1.1.20160309) [58] with the appropriate input dataset used as a control and a q-value cut-off of 0.01. This stringent threshold was selected to avoid overcalling peaks as a number of studies only had a single replicate for each condition. Insert size was estimated using the macs2 predicted function. For datasets with multiple replicates, only peaks which were at least partially present in at least two replicates were maintained in the dataset. Coverage counts were calculated as the number of datasets in which each position is covered by a peak using the BedTools intersect and merge functions [59].

### p53retriever pattern search algorithm

Sequences were identified by scanning the entire genome with a position weight matrix using the matchPWM function from the Biostrings package. The p53retriever pattern search algorithm was then utilized to score the putative p53 RE sequences from one (unlikely) to five (high) [27]. The algorithm was implemented in Python following the methodology described previously by Tebaldi et al., with slight modifications. First, it was optimised for parallelisation, in order to run efficiently genome-wide. Second, only full length p53 REs consisting of two half sites were considered and three-quarter sites and solo half sites were excluded. Lastly, the methodology used to select the position of the RE when the algorithm identifies two or more overlapping regions was altered. Specifically, REs overlapping by five or more base pairs were ranked first by score (highest to lowest), then by spacer length (shortest to longest), then by number of differences from the canonical p53 RE sequence (fewest to most), then the top RE in the overlapping region was selected. If all metrics were equal, the RE was selected arbitrarily.

Response elements falling within ChIP-Seq peaks were identified using the BedTools intersect function. The relationship between p53 retriever score and coverage was calculated using GAT [40]. GAT was run under the default settings with peaks at various coverage levels as intervals to intersect with the positions of the REs with each possible score from one to five. REs were categorised by coverage level as follows: if *C* is the median number of datasets (across the length of the RE) in which an RE was bound by p53, the REs were classified as unbound if *C* = 0, unlikely bound if 1 ≤ *C* < 3, lowly bound if 3 ≤ *C* < 5, moderately bound if 5 ≤ *C* < 10 and highly bound if 10 ≤ *C* < 27.

### Putative functional p53-REs

Polymorphism was established using the SNP data from phase 3 of the 1000 genomes project [60]. REs were considered to be polymorphic if they contained a SNP in 1000 Genomes in either half site (but not if the SNP was in the spacer region). cCREs regions (Registry V3) were downloaded from https://screen.encodeproject.org/ [1]. REs were considered to be cCREs if they reside within cCRE regions. The conservation score (PhyloP) calculated from the alignment of 100 vertebrate genomes per base was downloaded from the UCSC browser http://hgdownload.cse.ucsc.edu/goldenpath/hg38/phyloP100way/hg38.phyloP100way.bw. First we converted the 1-based p53-RE coordinates to 0-based, and calculated the average conservation scores for each REs using PhyloP scores and tool BigwigAverageOverBed. REs were attributed to genes if they reside within 10-Kb of genes annotated in Ensembl hg38 release 86.

### Genomics England 100K genomes project data

Whole-genome sequencing data from the 100K genomes project (100KGP) was supplied by Genomics England (GEL; Main Programme, release v14). All participants gave written, informed consent for their tissues to be sequenced as part of the 100KGP, and any sequencing data obtained from participants who subsequently withdrew their consent was excluded from our analyses. Participants’ DNA was sequenced by Illumina for GEL and was subject to quality control thresholds stipulated by GEL. Germline samples were required to be sequenced to yield at least 85 Gb of sequence data with a quality score of at least 30. Alignments provided by Illumina using their proprietary DRAGEN pipeline, were required to provide at least 95% coverage of the genome at 15X depth, or above. Individual gVCFs for the germline genomes of 78,195 participants were aggregated by Illumina for GEL to produce an aggregated gVCF (aggV2). File sizes were reduced, to facilitate easier handling and interrogation by bioinformatics tools, by dividing the aggV2 gVCF data into 1371 aggregated chunks stored in BCF format. The complete dataset and all analysis tools and applications were made available by GEL via their secure Research Environment.

### High-confidence sets of Pathogenic/Likely-Pathogenic and Benign/Likely-Benign variants from ClinVar

The entire ClinVar database (release date: 20/07/2020) was downloaded as a VCF file and was converted to a tab-separated file using the Genome Analysis Toolkit tool VariantsToTable (GATK 4.0.0.0, Broad Institute, MA, USA). To generate a set of pathogenic or likely pathogenic variants (P/LP), the ClinVar file was filtered using the grep command and search terms to capture records where the ‘CLNSIG’ field contained the annotations ‘Pathogenic’ or ‘Likely pathogenic’. Additional filtering of these variants was done to generate a high-confidence set, where the criteria in the ‘CLNREVSTAT fields were any of: ‘multiple submitters, no conflicts’, ‘reviewed by expert panel’ or ‘practice guideline’. Similarly, to generate a set of benign or likely benign variants (B/LB), the ClinVar file was filtered using the grep command and search terms to capture records where the ‘CLNSIG’ field contained the annotations ‘Benign’ or ‘Likely benign’. The ‘CLNREVSTAT’ criteria for inclusion in the high-confidence B/LB set were the same as for the P/LP set but with the addition of ‘criteria provided, single submitter’. This was to ensure that the high-confidence B/LB set was at least as large as the high-confidence P/LP set. We found that these slightly less stringent inclusion criteria were necessary to compensate for an inherent bias in ClinVar reflecting under reporting of benign variants relative to pathogenic variants.

### Selection of GEL cancer and control cohorts

To avoid the confounding effects of enrichment of variants in different ethnic populations, we decided to concentrate solely on European participants. Rather than relying on self-declared ethnicity, reported by participants, we used predicted ethnicities derived by GEL from principal component analysis of the aggregated aggV2 dataset. Ethnicities were resolved into five main categories: African, Admixed American, East Asian, European and South Asian. The probability threshold for each category was 0.8 and any participants scoring lower than this for all categories were listed as unassigned. Our cancer cohort consisted of the 88% of pan-cancer participants who were European (13,291 participants). For the control cohort we used European (78%) relatives of probands with rare diseases, which were not cancer predisposition syndromes (25, 086 participants). Diagnoses were obtained from GEL’s LabKey database.

### Extraction and enumeration of participants with P/LPs, B/LBs and p53-RE SNVs

Since the AggV2 aggregation provided by GEL had been done using genomic VCFs (gVCFs), which record loci in participants who do not have a variant at that position, as well as recording variants in those who do, we were able to enumerate the participants who had P/LPs, B/LBs and p53-RE SNVs and to calculate minor allele frequencies. We used the BCFtools [61] query command to extract records from AggV2 for each genomic location in the high-confidence P/LP and B/LB sets, as well as the list of p53-RE SNVs, for all participants in our cancer and control cohorts. Output from BCFtools was captured by Python (3.8.1) scripts which were run as parallel batch jobs on GEL’s IBM LSF high-performance cluster.

Additional filtering of records was done using Python scripts. Records where the quality control filter field was not ‘PASS’, were annotated as ‘Low quality’ and their genotypes were excluded from subsequent analysis. In accordance with established clinical guidelines [62], records where the variant allele frequency (VAF) was in the range 0.35 to 0.65 were defined as being heterozygous, whereas non-homozygous records outside of this range were considered mosaic and were annotated accordingly. Minor allele frequencies for each variant or SNV were calculated by counting alleles, with homozygous genotypes being counted as two and heterozygous genotypes being counted as one each for the major and minor alleles. For records of p53-RE SNVs, reports were generated by Python scripts which assigned SNVs according to whether they were present or not in cCREs and whether they were present in key bases or non-key bases.

### Statistical Analyses

All statistical tests, including Fisher’s exact test for enrichment, were done using R (4.1.2; The R foundation for statistical computing) with appropriate correction of p-values for multiple hypothesis testing.

## Data Availability

All datasets used in this study are publicly available, with details included in the manuscript and supplementary information. All data produced in the present study are available upon request.

## Acknowledgements

Funding for GLB for this work was provided by the Ludwig Institute for Cancer Research and University of Birmingham. PZ was funded in part by the Ludwig Institue for Cancer Research. DB was funded by the University of Birmingham. This research was made possible through access to the data and findings generated by the 100,000 Genomes Project. The 100,000 Genomes Project is managed by Genomics England Limited (a wholly owned company of the Department of Health). The 100,000 Genomes Project is funded by the National Institute for Health Research and NHS England. The Wellcome Trust, Cancer Research UK and the Medical Research Council have also funded research infrastructure. The 100,000 Genomes Project uses data provided by patients and collected by the National Health Service as part of their care and support. Funding from Sarcoma UK (SUKG01.2018), the Tom Prince Cancer Trust and the Bone Cancer Research Trust contributed to the collection and processing of the samples. AMF and NP are supported by the National Institute for Health Research, UCLH Biomedical Research Centre and the UCL Experimental Cancer Centre. SDN holds a Jean Shanks Foundation - Pathological Society Clinical PhD Fellowship. PVL was supported by the Francis Crick Institute, which receives its core funding from Cancer Research UK (FC001202), the UK Medical Research Council (FC001202), and the Wellcome Trust (FC001202). For the purpose of open access, the authors have applied a CC BY public copyright licence to any author accepted manuscript version arising from this submission. P.V.L. is a Winton Group Leader in recognition of the Winton Charitable Foundation’s support towards the establishment of The Francis Crick Institute.

P.V.L. is a CPRIT Scholar in Cancer Research and acknowledges CPRIT grant support (RR210006).

