## Supplementary Table 6 for "A novel map of human p53 response elements uncovers evidence of selection pressures and variants similar to Li-Fraumeni Syndrome mutations"

**Supplementary Table 6: Known pathogenic or likely pathogenic variants in TP53 associated with Li-Fraumeni syndrome from ClinVar.**

| Genomic location | Amino Acid change | Consequence | dbSNP rs ID | AF_ESP | AF_EXAC | AF_TGP |
|---|---|---|---|---|---|---|
| g.7669692T>C | NA | splice_acceptor_variant | rs587781664 | NA | NA | NA |
| g.7670669G>T | NA | missense_variant, 3_prime_UTR_variant | rs397516434 | NA | NA | NA |
| g.7670678A>G | p.Leu344Pro | missense_variant, 3_prime_UTR_variant | rs121912662 | NA | NA | NA |
| g.7670684C>G | p.Arg342Pro | missense_variant, 3_prime_UTR_variant | rs375338359 | NA | NA | NA |
| g.7670686del | NA | frameshift_variant, 3_prime_UTR_variant | rs1131691022 | NA | NA | NA |
| g.7670685G>A | NA | nonsense, 3_prime_UTR_variant | rs730882029 | NA | NA | NA |
| g.7670699C>T | p.Arg337His | missense_variant, 3_prime_UTR_variant | rs121912664 | NA | 0.00001 | NA |
| g.7670700G>A | p.Arg337Cys | missense_variant, 3_prime_UTR_variant | rs587782529 | NA | 0.00001 | NA |
| g.7670716C>G | NA | splice_acceptor_variant | rs587782272 | NA | NA | NA |
| g.7673535del | NA | splice_donor_variant | rs1131691033 | NA | NA | NA |
| g.7673534C>G | NA | splice_donor_variant | rs11575997 | NA | NA | NA |
| g.7673535C>T | NA | synonymous_variant | rs11575996 | NA | NA | NA |
| g.7673552C>A | NA | nonsense | rs876659384 | NA | NA | NA |
| g.7673579G>A | NA | nonsense | rs764735889 | NA | NA | NA |
| g.7673609C>T | NA | splice_acceptor_variant | rs587781702 | NA | NA | NA |
| g.7673610T>C | NA | splice_acceptor_variant | rs397516439 | NA | NA | NA |
| g.7673699A>C | NA | splice_donor_variant | rs1131691016 | NA | NA | NA |
| g.7673700C>T | NA | splice_donor_variant | rs1131691039 | NA | NA | NA |
| g.7673704G>A | NA | nonsense | rs121913344 | NA | NA | NA |
| g.7673728C>A | NA | nonsense | rs201744589 | NA | NA | NA |
| g.7673764C>T | p.Glu286Lys | missense_variant | rs786201059 | NA | NA | NA |
| g.7673767C>T | NA | missense_variant | rs112431538 | NA | NA | NA |
| g.7673775C>G | NA | missense_variant | rs730882008 | NA | NA | NA |
| g.7673776G>A | p.Arg282Trp | missense_variant | rs28934574 | NA | 0.00002 | NA |
| g.7673778T>A | p.Asp281Val | missense_variant | rs587781525 | NA | NA | NA |
| g.7673778T>C | p.Asp281Gly | missense_variant | rs587781525 | NA | NA | NA |
| g.7673796C>T | p.Cys275Tyr | missense_variant | rs863224451 | NA | NA | NA |
| g.7673802C>A | NA | missense_variant | rs28934576 | NA | NA | NA |
| g.7673802C>G | p.Arg273Pro | missense_variant | rs28934576 | NA | NA | NA |
| g.7673802C>T | p.Arg273His | missense_variant | rs28934576 | NA | NA | 0.0002 |
| g.7673803G>A | p.Arg273Cys | missense_variant | rs121913343 | NA | NA | NA |
| g.7673803G>C | NA | missense_variant | rs121913343 | NA | NA | NA |

| | | | | | | |
|---|---|---|---|---|---|---|
| g.7673803G>T | NA | missense_variant | rs121913343 | NA | NA | NA |
| g.7673811A>G | NA | missense_variant | rs1057519986 | NA | NA | NA |
| g.7673820dup | NA | frameshift_variant | rs1597362206 | NA | NA | NA |
| g.7673821G>A | p.Arg267Trp | missense_variant | rs55832599 | NA | NA | NA |
| g.7673826A>G | p.Leu265Pro | missense_variant | rs879253942 | NA | NA | NA |
| g.7673830del | NA | frameshift_variant | rs1060501194 | NA | NA | NA |
| g.7673838C>T | NA | splice_acceptor_variant | rs1555525367 | NA | NA | NA |
| g.7674191C>T | p.Glu258Lys | missense_variant | rs121912652 | NA | 0.00001 | NA |
| g.7674199_7674201ATG[1] | NA | inframe_deletion | rs1064794309 | NA | NA | NA |
| g.7674220C>A | p.Arg248Leu | missense_variant | rs11540652 | NA | NA | NA |
| g.7674220C>T | p.Arg248Gln | missense_variant | rs11540652 | NA | 0.00006 | NA |
| g.7674221G>A | p.Arg248Trp | missense_variant | rs121912651 | NA | 0.00001 | NA |
| g.7674226A>T | NA | missense_variant | rs587780074 | NA | NA | NA |
| g.7674227T>C | p.Met246Val | missense_variant | rs483352695 | NA | NA | NA |
| g.7674229C>A | NA | missense_variant | rs121912656 | NA | 0.00001 | NA |
| g.7674229C>T | p.Gly245Asp | missense_variant | rs121912656 | NA | NA | NA |
| g.7674230C>A | p.Gly245Cys | missense_variant | rs28934575 | NA | NA | NA |
| g.7674230C>T | p.Gly245Ser | missense_variant | rs28934575 | NA | 0.00001 | NA |
| g.7674232C>T | NA | missense_variant | rs985033810 | NA | NA | NA |
| g.7674233C>T | NA | missense_variant | rs1057519989 | NA | NA | NA |
| g.7674238C>T | p.Cys242Tyr | missense_variant | rs121912655 | NA | NA | NA |
| g.7674241G>A | p.Ser241Phe | missense_variant | rs28934573 | NA | NA | NA |
| g.7674241G>C | p.Ser241Cys | missense_variant | rs28934573 | NA | 0.00001 | NA |
| g.7674250C>T | p.Cys238Tyr | missense_variant | rs730882005 | NA | 0.00002 | NA |
| g.7674251A>G | NA | missense_variant | rs1057519981 | NA | NA | NA |
| g.7674262T>C | p.Tyr234Cys | missense_variant | rs587780073 | NA | 0.00001 | NA |
| g.7674263A>G | NA | missense_variant | rs864622237 | NA | NA | NA |
| g.7674291C>T | NA | splice_acceptor_variant | rs878854073 | NA | NA | NA |
| g.7674859C>T | NA | synonymous_variant | rs267605076 | NA | NA | NA |
| g.7674869del | NA | frameshift_variant | rs878854071 | NA | NA | NA |
| g.7674872T>C | p.Tyr220Cys | missense_variant | rs121912666 | NA | 0.00003 | NA |
| g.7674890T>C | NA | missense_variant | rs1057519992 | NA | NA | NA |
| g.7674893C>T | p.Arg213Gln | missense_variant | rs587778720 | NA | 0.00001 | NA |
| g.7674894G>A | NA | nonsense | rs397516436 | NA | 0.00001 | NA |
| g.7674905_7674906del | NA | frameshift_variant | rs1057517840 | NA | NA | NA |
| g.7674945G>A | NA | nonsense | rs397516435 | NA | 0.00001 | NA |
| g.7674947A>G | p.Ile195Thr | missense_variant | rs760043106 | NA | 0 | NA |
| g.7674953T>C | p.His193Arg | missense_variant | rs786201838 | NA | NA | NA |
| g.7674953T>G | NA | missense_variant | rs786201838 | NA | NA | NA |

| | | | | | | |
|---|---|---|---|---|---|---|
| g.7674954G>A | p.His193Tyr | missense_variant | rs876658468 | NA | NA | NA |
| g.7674957G>A | NA | nonsense | rs866380588 | NA | NA | NA |
| g.7675052C>A | NA | splice_donor_variant | rs1131691042 | NA | NA | NA |
| g.7675052C>T | NA | splice_donor_variant | rs1131691042 | NA | NA | NA |
| g.7675070C>G | NA | missense_variant | rs397514495 | NA | NA | NA |
| g.7675070C>T | p.Arg181His | missense_variant | rs397514495 | NA | 0.00002 | NA |
| g.7675074C>T | p.Glu180Lys | missense_variant | rs879253911 | NA | NA | NA |
| g.7675075A>T | NA | missense_variant | rs876660821 | NA | NA | NA |
| g.7675077G>A | p.His179Tyr | missense_variant | rs587780070 | NA | NA | NA |
| g.7675084del | NA | frameshift_variant | rs786202525 | NA | NA | NA |
| g.7675080G>C | NA | missense_variant | rs1064795203 | NA | NA | NA |
| g.7675082G>C | NA | missense_variant | rs751477326 | NA | 0.00002 | NA |
| g.7675085C>T | p.Cys176Tyr | missense_variant | rs786202962 | NA | NA | NA |
| g.7675088C>A | p.Arg175Leu | missense_variant | rs28934578 | NA | NA | NA |
| g.7675088C>T | p.Arg175His | missense_variant | rs28934578 | NA | 0.00001 | NA |
| g.7675089G>C | NA | missense_variant | rs138729528 | NA | NA | NA |
| g.7675094A>G | NA | missense_variant | rs1057519747 | NA | NA | NA |
| g.7675095C>T | p.Val173Met | missense_variant | rs876660754 | NA | NA | NA |
| g.7675102_7675105del | NA | frameshift_variant | rs1555526082 | NA | NA | NA |
| g.7675113G>A | NA | nonsense | rs1555526097 | NA | NA | NA |
| g.7675119G>A | NA | nonsense | rs730882001 | NA | NA | NA |
| g.7675124T>C | p.Tyr163Cys | missense_variant | rs148924904 | NA | NA | NA |
| g.7675138_7675139delinsAA | NA | missense_variant, 5_prime_UTR_variant | rs1567553501 | NA | NA | NA |
| g.7675139C>A | NA | missense_variant, 5_prime_UTR_variant | rs587782144 | NA | NA | NA |
| g.7675139C>G | p.Arg158Pro | missense_variant, 5_prime_UTR_variant | rs587782144 | NA | NA | NA |
| g.7675139C>T | p.Arg158His | missense_variant, 5_prime_UTR_variant | rs587782144 | NA | 0.00001 | NA |
| g.7675161dup | NA | frameshift_variant, 5_prime_UTR_variant | rs730882019 | NA | NA | NA |
| g.7675157G>A | p.Pro152Leu | missense_variant, 5_prime_UTR_variant | rs587782705 | NA | 0.00003 | NA |
| g.7675161G>A | p.Pro151Ser | missense_variant, 5_prime_UTR_variant | rs28934874 | NA | NA | NA |
| g.7675161G>T | p.Pro151Thr | missense_variant, 5_prime_UTR_variant | rs28934874 | NA | NA | NA |
| g.7675174C>T | NA | nonsense, 5_prime_UTR_variant | rs1131691026 | NA | NA | NA |
| g.7675175C>T | NA | nonsense, 5_prime_UTR_variant | rs1206165503 | NA | NA | NA |
| g.7675184A>G | NA | missense_variant, 5_prime_UTR_variant | rs1555526241 | NA | NA | NA |
| g.7675190C>T | p.Cys141Tyr | missense_variant, | rs587781288 | NA | NA | NA |

| | | | | | | |
|---|---|---|---|---|---|---|
| | | 5_prime_UTR_variant | | | | |
| g.7675209A>C | NA | missense_variant, 5_prime_UTR_variant | rs1057519975 | NA | NA | NA |
| g.7675220T>A | NA | missense_variant, 5_prime_UTR_variant | rs1131691037 | NA | NA | NA |
| g.7675237C>T | NA | splice_acceptor_variant, 5_prime_UTR_variant | rs868137297 | NA | NA | NA |
| g.7675238T>C | NA | splice_acceptor_variant, 5_prime_UTR_variant | rs786202799 | NA | NA | NA |
| g.7675994dup | NA | NA | rs1555526470 | NA | NA | NA |
| g.7675992A>G | NA | splice_donor_variant | rs1555526469 | NA | NA | NA |
| g.7675992A>T | NA | splice_donor_variant | rs1555526469 | NA | NA | NA |
| g.7675993C>A | NA | splice_donor_variant | rs1567555445 | NA | NA | NA |
| g.7675994C>T | NA | synonymous_variant | rs55863639 | NA | NA | NA |
| g.7675995G>C | NA | missense_variant | rs786201057 | NA | NA | NA |
| g.7675997G>T | NA | nonsense | rs1555526478 | NA | NA | NA |
| g.7676003_7676004CA[1] | NA | frameshift_variant | rs587780067 | NA | NA | NA |
| g.7676042_7676048dup | NA | frameshift_variant | rs1131691004 | NA | NA | NA |
| g.7676040C>A | NA | missense_variant | rs11540654 | NA | NA | NA |
| g.7676040C>G | p.Arg110Pro | missense_variant | rs11540654 | NA | NA | NA |
| g.7676041_7676042del | NA | frameshift_variant | rs1064795434 | NA | NA | NA |
| g.7676060G>C | NA | nonsense | rs1597373901 | NA | NA | NA |
| g.7676098_7676120del | NA | frameshift_variant | rs886041861 | NA | NA | NA |
| g.7676096C>T | NA | nonsense | rs876660548 | NA | NA | NA |
| g.7676098dup | NA | frameshift_variant | rs1597374152 | NA | NA | NA |
| g.7676158dup | NA | frameshift_variant | rs730882018 | NA | NA | NA |
| g.7676210C>T | NA | nonsense | rs1064794618 | NA | NA | NA |
| g.7676211C>T | NA | nonsense | rs876658483 | NA | NA | NA |
| g.7676273C>T | NA | splice_acceptor_variant | rs1597375294 | NA | NA | NA |
| g.7676381C>A | NA | splice_donor_variant | rs1131691003 | NA | NA | NA |

Variants were obtained from the high-confidence set of ClinVar P/LPs (Materials and Methods) by using the search terms ‘Li-Fraumeni’ and ‘TP53:7157’. Abbreviations: AF = Allelic frequency, ESP = Exome Sequencing Project database,  EXAC = Exome Aggregation Consortium database, TGP = 1000 Genomes Project database, NA = not available.
