## Supplementary Table 7 for "A novel map of human p53 response elements uncovers evidence of selection pressures and variants similar to Li-Fraumeni Syndrome mutations"

**Supplementary Table 7: NHS Hospitals and regions from which Genomics England participants were recruited.**

| Recruiting Hospital | Region |
|---|---|
| Norfolk and Norwich University Hospitals NHS Foundation Trust | East of England |
| Queen’s Medical Centre University Hospital Pharmacy Production Unit | East of England |
| University Hospitals of Leicester NHS Trust | East of England |
| Salford Royal Hospital | Greater Manchester |
| University Hospital of South Manchester NHS Foundation Trust | Greater Manchester |
| Whipps Cross University Hospital TSDU | North Thames |
| Royal National Orthopaedic Hospital NHS Trust (Stanmore) | North Thames |
| University College London Hospital | North Thames |
| Aintree University Hospital | North West |
| Royal Orthopaedic Hospital | West Midlands |
| North Bristol NHS Trust Sarcoma Cancer and Breast Cancer Clinics | West of England |
